# Public opinion on a mandatory COVID-19 vaccination policy in France: a cross sectional survey

**DOI:** 10.1101/2021.07.05.21260017

**Authors:** Amandine Gagneux-Brunon, Elisabeth Botelho-Nevers, Marion Bonneton, Patrick Peretti-Watel, Pierre Verger, Odile Launay, Jeremy K. Ward

## Abstract

**Objectives:** Reaching the last pockets of unvaccinated people is challenging, and has led to consider COVID-19 mandatory vaccination. Our aim was to assess attitudes toward COVID-19 mandatory vaccination in France before the announcement and factors associated with opposition to this type of policy.

**Methods:** Between the 10th and the 23rd of May 2021, we conducted a cross-sectional online survey among a representative sample of the French population aged 18 and over and a specific sample of the French Senior Population over 65.

**Results:** Among 3,056 respondents, 1,314 (43.0 %) were in favor of mandatory COVID-19 vaccination, 1,281 (41.9 %) were opposed to such a policy, and 461 (15.1 %) were undecided. Among opponents to COVID-19 mandatory vaccination for the general population, 385 (30.05 %) were in favor of a mandatory COVID-19 vaccination for healthcare workers (HCWs). In multivariate analysis, age groups 18-24 years, and 25-34 years were significantly more opposed than the reference group (>75 years old) with respective adjusted odds ratio (aOR) and 95 % confidence interval (95 % CI) 4.67 (1.73-12.61) and 3.74 (1.57-8.93). No intention of getting COVID-19 vaccine was strongly associated with opposition to mandatory vaccination with aOR 10.67 (95 % CI 6.41-17.76). In comparison with partisans of the center, partisans of the far left and green parties were more likely to be opposed to COVID-19 mandatory vaccine with respective aOR (95 % CI) 1;89 (1.06-3.38) and 2.08 (1.14-3.81).

**Conclusion:** Attitudes toward mandatory COVID-19 vaccination are split in the French general population, and the debate might become politicized.

## Introduction

COVID-19 has been responsible for more than 225 million cases and more than 4.6 million deaths worldwide up to the 14^th^ of September 2021 [1]. Vaccines were developed at “a pandemic speed”[2]. More than 5 billion COVID-19 vaccines had been administered, worldwide, by the end of August 2021 [3]. After 5 to 6 months of covid-19 vaccination campaigns, many high income countries have reached their coverage plateau (60 % of the entire population) [3]. In France, all individuals over 12 years of age have been eligible for COVID-19 vaccination since the 1^st^ of June 2021. On the July 1^st^ 2021, 51. % of the French general population (59.8 % of the eligible population) had received a first dose of COVID-19 [4].

As has been seen in the past with childhood immunizations, vaccinating a majority of the population is easier than reaching the last pockets of unvaccinated people, the unwilling or weakly motivated [5,6]. Faced with this challenge in the past, many countries have resorted to various forms of vaccine mandates [7].While recourse to constraint in its various forms can be effective in raising vaccine coverage –particularly by pushing those who wait, those who refuse, to act – it also presents the risk of antagonizing part of the public, causing reactance and stimulating antivaccine movements [8]. Because the debate around mandatory COVID vaccination is emerging in many countries, it is crucial to understand the conditions in which this policy can be widely accepted. France was among the most vaccine-hesitant countries in the world before the epidemic of COVID [9,10] and hesitancy toward covid-19 vaccination has remained higher than in most neighboring countries throughout the period [11]. Studying attitudes towards vaccine mandates in such a context can help highlight the variety of factors influencing the acceptability of coercive measures including preferences for political parties [12], identify target groups, and develop specific interventions to reduce reactance. Although HCWs were identified as a priority target group for COVID-19 vaccination, only 60 % of the French HCWs had received a dose of COVID-19 vaccine on the 1^st^ of July [4]. To increase COVID-19 vaccine coverage in HCWs, COVID-19 vaccine mandates appeared as a solution. In France, the compulsory vaccination against hepatitis B led to a significantly increase in vaccination coverage and reduced the differences between professional categories [13].

In a survey carried out in May 2021, participants were asked for their opinion about COVID-19 vaccine mandates for the general population and HCWs [14]. In this context, it seems interesting to assess opinions about mandatory vaccination prior to the implementation, and to identify factors associated with opposition to COVID-19 vaccine mandates in France.

## Methods

### Design and sample

Between the 10^th^ and the 23^rd^ of May 2021, we conducted a cross-sectional online survey among a sample of the French population aged 18 and over, with participants who were randomly selected from an existing online research panel of more than 750,000 nationally representative households of the general population (Bilendi SA^®^). A quota sampling method was applied to achieve a sample of 1,514 respondents, representative for the French adult population in terms of age, gender, occupation and population in the area of residence. A total of 50,200 invitations were sent to reach this sample (Response rate 3.1 %). An additional sample of 1,544 French residents 65 years of age and older selected from the same panel, representative of the general “senior” population in terms of gender and age was added because the survey also aimed to identify reasons for non-vaccination in the elderly. A total of 5,700 additional persons over 65 years of age were invited to answer the survey to obtain this extra sample (Response rate 27.1 %). Prior information on the panelists was used to determine eligibility and to select a stratified random sample with oversampling of panelists over 65 years of age. To limit coverage bias, due to the fact that not all people use the internet, and, among users, that not all of them are willing to participate in web surveys, random sampling was stratified to match French official census statistics for gender, age, occupation (8 categories), population in the area of residence (5 categories) and region (12 categories). In addition, a survey weight that takes into account gender, age, region and size of residence area was calculated and assigned to each response. The study design was approved by the ethical committee of the University Hospital Institute Méditerranée Infection (#2021–001).

### Data collected

In addition to background socio-economic variables (gender, age, profession), we collected intention or history of COVID-19 vaccination, concerns about COVID-19 and opinion of vaccines in general. Respondents were asked to which French political party they felt the closest (among a quite comprehensive list of 17 parties), and responses were encoded into: Far-Left, Green party, Left, Center, Right governmental parties, Far-Right, and feeling close to no party. Regarding COVID-19 mandatory vaccination, respondents were asked whether they think that vaccination against COVID should be mandatory for the entire population, the question was “Do you think that COVID-19 vaccination should be mandatory for all?”. Respondents against mandatory COVID-19 vaccination were asked whether they think that COVID-19 vaccination should be mandatory for HCWs “Do you think that COVID-19 vaccination should be mandatory for HCWs?”. For both questions, the answers were yes, no or don’t know.

### Statistical analysis

Attitude toward a mandatory COVID-19 vaccination for the general population were merged into a binary outcome: ‘opposition to COVID-19 mandatory vaccine policy’ equaled 1 if participants answered No, otherwise the value was 0. We chose this dichotomization, while we considered that undecided individuals will not be those who will strongly express their opposition. We first used bivariate analyses, and chi-square tests in cross-tabulations and a bivariate logistic regression to investigate factors associated with opposition to mandatory COVID-19 vaccine, using respondents’ socio-economic background, COVID-19 concern, and political preferences as covariates. In a second part, we aimed to better describe the population of individuals reluctant to COVID-19 vaccine mandates for all, but in favor of mandatory COVID-19 vaccination for HCWs. In the regression model, we used bivariate analyses and a bivariate logistic regression to investigate factors associated with the attitude toward a COVID-19 vaccine mandate for HCWs in respondents opposed to a mandatory COVID-19 vaccine policy for all. In regression models, we used a forward stepwise selection method (entry threshold p <0.2) to retain statistically significant covariates only.

## Results

A total of 3,056 individuals answered the questionnaire (1,455 men, 47.6 %). Among the respondents, 1,314 (43.0 %) were in favor of mandatory COVID-19 vaccination, 1,281 (41.9 %) were opposed to such a policy, and 461 (15.1 %) were undecided (Table 1).

**Table 1.** Comparison between respondents with favorable or undecided opinion and respondents with negative opinion towards COVID-19 vaccine mandates (n: number, CEO: chief executive officer).

|  | Favorable or undecided opinion toward COVID-19 vaccination mandates (n=1,775) |  | Opposition toward COVID-19 vaccination mandates (n=1,281) |  | p value |
| --- | --- | --- | --- | --- | --- |
|  | n | % | n | % |  |
| Gender |  |  |  |  |  |
| Male (n=1,455) | 850 | 58.4 | 605 | 41.6 | 0.83 |
| Female (n=1,601) | 925 | 57.8 | 676 | 42.2 |  |
| Age groups |  |  |  |  |  |
| 18-24 (n=315) | 146 | 46.5 | 169 | 53.51 | <0.005 |
| 25-34 (n=447) | 173 | 38.6 | 274 | 61.37 |  |
| 35-49 (n=737) | 373 | 50.6 | 364 | 49.36 |  |
| 50-64 (n=745) | 444 | 59.6 | 301 | 40.37 |  |
| 65-74 (n=439) | 334 | 75.9 | 105 | 24.02 |  |
| 75 and older (n=373) | 305 | 81.8 | 68 | 18.17 |  |
| Socio-professional categories |  |  |  |  | <0.005 |
| Farmers, Artisans, shopkeepers. CEOs (n=130) | 62 | 47.8 | 68 | 52.2 |  |
| Executives and intellectual professionals (n=327) | 173 | 53.1 | 154 | 46.9 |  |
| Intermediate professions (n=423) | 223 | 52.7 | 200 | 47.2 |  |
| Employees (n=437) | 213 | 48.7 | 224 | 51.3 |  |
| Blue collar Workers (n=336) | 140 | 41.7 | 196 | 58.3 |  |
| Retired (n=1,032) | 768 | 74.5 | 264 | 25.5 |  |
| Unemployed (n=371) | 196 | 52.7 | 175 | 47.3 |  |
| Healthcare workers |  |  |  |  | 0.16 |
| Yes (n=291) | 151 | 51.9 | 140 | 48.1 |  |
| No (n= 2765) | 1624 | 58.7 | 1141 | 41.3 |  |
| Fear of getting infected |  |  |  |  | <0.005 |
| Not frightened (n=899) | 438 | 48.8 | 461 | 51.2 |  |
| Not very frightened (n=809) | 485 | 60.1 | 323 | 39.9 |  |
| Very frightened (n=778) | 456 | 58.6 | 322 | 41.4 |  |
| Extensively frightened (n=571) | 396 | 69.3 | 175 | 30.7 |  |
| Intention of getting vaccinated |  |  |  |  | <0.005 |
| Yes or already vaccinated (n=2,348) | 1670 | 71.1 | 678 | 28.9 |  |
| No (n=708) | 105 | 14.8 | 603 | 85.2 |  |
| Trust in pharmaceutical companies |  |  |  |  | <0.005 |
| Yes (n = 1,317) | 922 | 70.0 | 395 | 30.0 |  |
| no (n = 1,739) | 853 | 49.1 | 886 | 50.9 |  |
| Trust in government during the pandemic |  |  |  |  | <0.005 |
| Yes (n = 1,428) | 1037 | 72.6 | 391 | 27.4 |  |
| no (n = 1,628) | 738 | 45.4 | 890 | 54.6 |  |
| Favorable opinion toward vaccines in general |  |  |  |  | <0.005 |
| Yes (n = 2,349) | 1561 | 66.5 | 788 | 33.5 |  |
| No/undecided (n = 707) | 214 | 30.3 | 493 | 69.7 |  |
| Partisan preference |  |  |  |  | <0.005 |
| Others (n = 97) | 59 | 61.3 | 38 | 38.7 |  |
| No declared preference (n=1,027) | 557 | 54.2 | 471 | 45.8 |  |
| Far left (n=252) | 107 | 42.3 | 145 | 57.7 |  |
| Left (n=272) | 174 | 63.8 | 98 | 36.2 |  |
| Green party (n=217) | 104 | 47.9 | 113 | 52.1 |  |
| Center (n=449) | 337 | 75.0 | 112 | 25.0 |  |
| Right (n=292) | 209 | 71.5 | 83 | 28.5 |  |
| Far right (n=450) | 229 | 51.0 | 221 | 49.0 |  |
| Science does people (n=2,756) |  |  |  |  | <0.005 |
| more good than harm (n = 1,131) | 713 | 64.2 | 418 | 35.8 |  |
| about as much good as harm (n = 1,448) | 794 | 54.8 | 654 | 45.2 |  |
| more harm than good (n = 177) | 52 | 29.6 | 125 | 70.4 |  |

All variables with a p value <0.1 in univariate analysis were integrated in the regression model. aOR: adjusted odds ratio. Variables associated with the attitude toward a mandatory COVID-19 vaccination policy are in Bold (with p value <0.05)

**Table 2:**
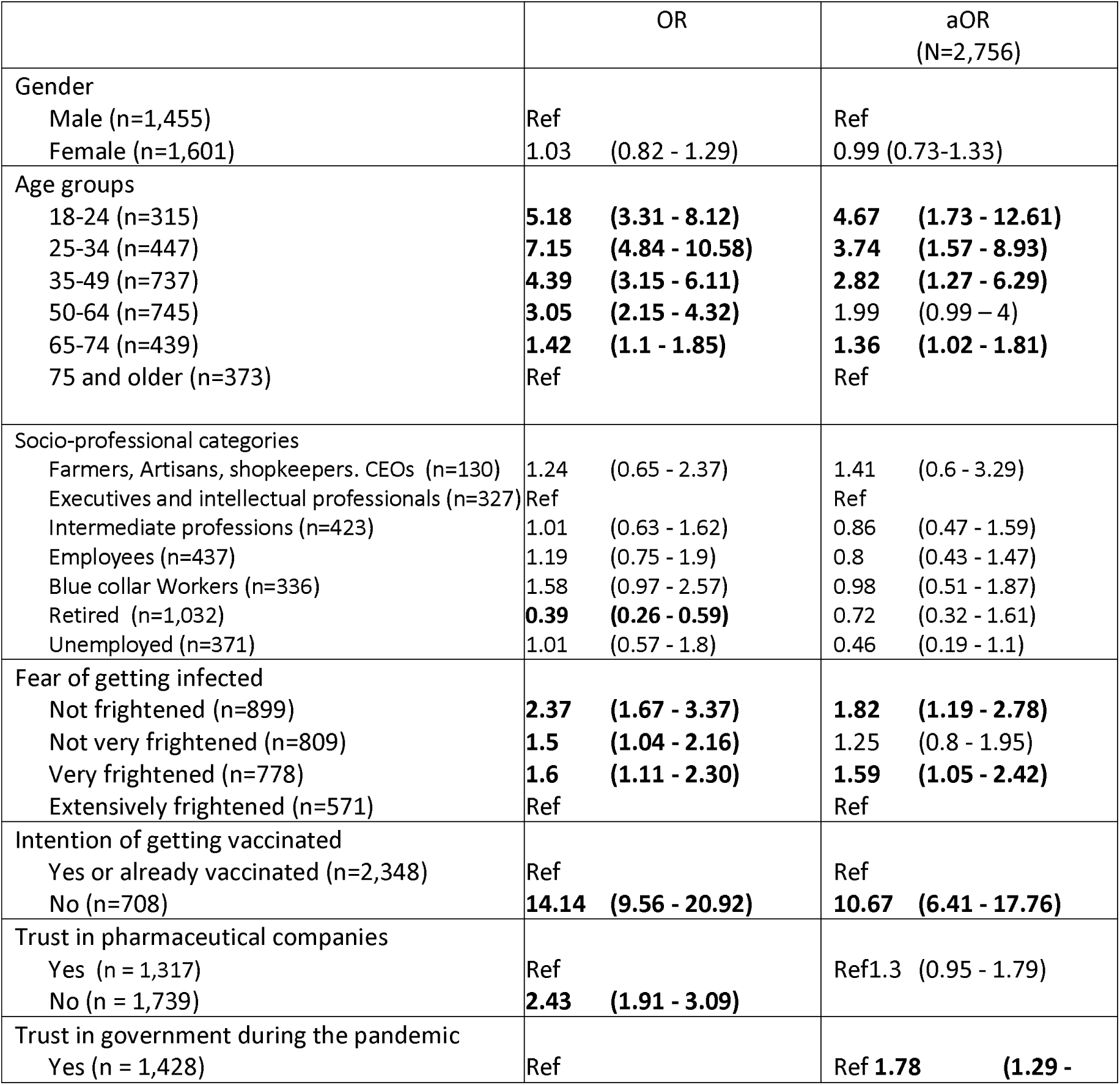
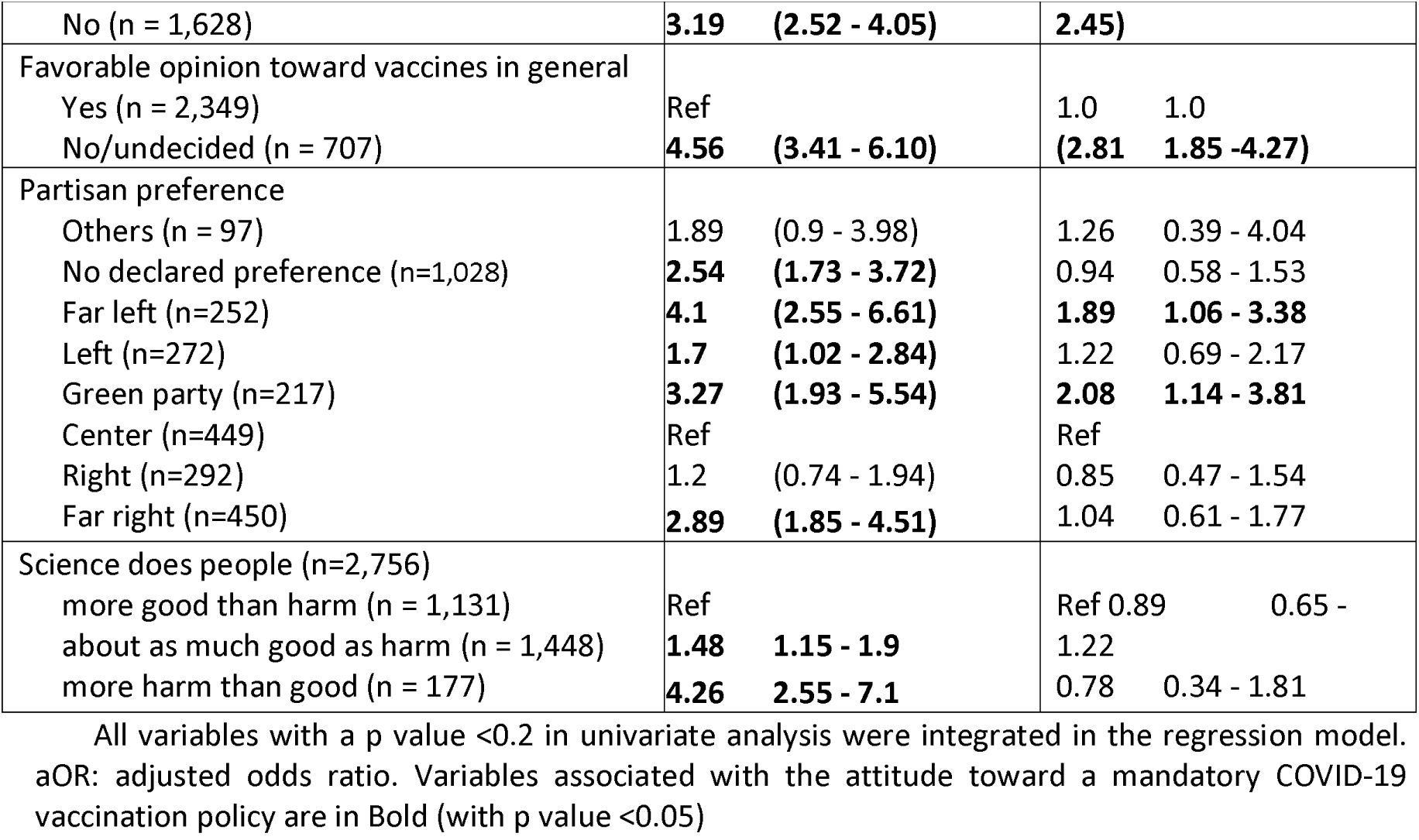
Factors associated with opposition to COVID-19 mandates for the general population. (Ref: reference, variables with a p-value <0.05 are in bold, OR: odds ratio, aOR: adjusted odds ratio, (n: number, CEO: chief executive officer)

Opinion differed between age groups, 61.4 % of the respondents aged from 25 to 34 years were opposed to COVID-19 in contrast with 18.2 % of individuals over the age of 75Among the respondents who intended to get vaccinated or had already been vaccinated, 28.9 % were opposed to mandatory COVID-19 vaccination.

The multivariate analysis confirmed that opinion toward a mandatory COVID-19 policy differed between age groups; younger individuals were more likely to be opposed to a mandatory COVID-19 vaccination (Table2). COVID-19 vaccine personal refusal was an important predictor of opposition to a mandatory COVID-19 vaccination with an aOR 10.67 (95 % CI 6.41 - 17.76). Differences in attitude to a COVID-19 mandatory vaccination were observed depending on political affiliation. Low trust in the government was also associated with reluctance to accept a mandatory COVID-19 vaccine policy with aOR 1.78 (95 % CI°1.29 - 2.45). Respondents with an unfavorable or no opinion about vaccination in general were also reluctant to accept a mandatory COVID-19 vaccine policy with an aOR 2.81 (95 % CI 1.85 -4.27).

Among the 1,281 individuals opposed to mandatory COVID-19 vaccination, 386 (30.1 %) were nevertheless in favor of a mandatory COVID-19 vaccination for HCWs. Individuals against a mandatory COVID-19 vaccine policy but accepting mandatory vaccine for HCWs represented 12.6 % of the sample. Factors associated with acceptance of a mandatory COVID-19 vaccine policy limited to HCWs are depicted in Table 3 and Table 4..

**Table 3:** Factors associated with acceptance of a mandatory COVID-19 vaccine policy only for Healthcare workers in the French general population in opponents to a COVID-19 mandatory vaccine policy for the general population (n=1,281) (n: number, CEO: chief executive officer)

|  | Negative or undecided opinion toward COVID-19 vaccine mandates for HCWs n=895 |  | Favorable opinion toward COVID-19 vaccine mandates for HCWs n= 386 |  | p value |
| --- | --- | --- | --- | --- | --- |
|  | n | % | n | % |  |
| Gender |  |  |  |  | 0.4 |
| Male (n=605) | 411 | 68.0 | 194 | 32.0 |  |
| Female (n=676) | 484 | 71.6 | 192 | 28.4 |  |
| Age groups |  |  |  |  | <0.005 |
| 18-24 (n=168) | 114 | 67.9 | 54 | 32.1 |  |
| 25-34 (n=275) | 218 | 79.3 | 57 | 20.7 |  |
| 35-49 (n=367) | 282 | 76.7 | 85 | 23.3 |  |
| 50-64 (n=301) | 189 | 62.7 | 112 | 37.3 |  |
| 65-74 (n=105) | 62 | 59.5 | 43 | 40.5 |  |
| 75 and older (n=68) | 33 | 48.3 | 35 | 51.7 |  |
| Socio-professional categories |  |  |  |  | 0.11 |
| Farmers, Artisans, shopkeepers, CEOs(n=68) | 53 | 77.9 | 15 | 22.1 |  |
| Executives and intellectual professionals (n=154) | 105 | 68.2 | 49 | 37.8 |  |
| Intermediate professions (n=200) | 142 | 71.0 | 58 | 29.0 |  |
| Employees (n=224) | 173 | 77.2 | 51 | 22.8 |  |
| Workers (n=196) | 145 | 73.9 | 51 | 26.1 |  |
| Retired (n=263) | 152 | 57.8 | 111 | 42.2 |  |
| Unemployed (n=175) | 125 | 71.4 | 50 | 28.6 |  |
| Healthcare workers |  |  |  |  | 0.54 |
| Yes (n=140) | 103 | 73.6 | 37 | 26.4 |  |
| No (n=1141) | 792 | 69.4 | 349 | 30.6 |  |
| Fear of getting infected |  |  |  |  | 0.09 |
| Not frightened (n=461) | 344 | 74.6 | 117 | 25.4 |  |
| A little frightened (n=323) | 237 | 73.4 | 86 | 26.6 |  |
| Very frightened (n=222) | 145 | 65.3 | 77 | 34.6 |  |
| Extremely frightened (n=175) | 105 | 60.0 | 70 | 40.0 |  |
| Intention of getting vaccinated |  |  |  |  | <0.005 |
| Yes or already vaccinated (n=678) | 389 | 57.4 | 289 | 42.6 |  |
| No (n=603) | 506 | 83.9 | 97 | 16.1 |  |
| Trust in pharmaceutical companies |  |  |  |  | <0.005 |
| Yes (n = 395) | 239 | 60.6 | 156 | 39.4 |  |
| No (n = 886) | 656 | 74.1 | 230 | 25.9 |  |
| Trust in government during the pandemic |  |  |  |  | <0.005 |
| Yes (n = 391) | 238 | 60.9 | 153 | 39.1 |  |
| No (n = 890) | 658 | 73.9 | 233 | 26.1 |  |
| Favorable opinion toward vaccines in general |  |  |  |  | 0.46 |
| Yes (n = 789) | 542 | 68.7 | 247 | 31.3 |  |
| No/undecided (n = 493) | 354 | 71.8 | 139 | 28.2 |  |
| Partisan preference |  |  |  |  | 0.08 |
| Others (n = 37) | 29 | 78.3 | 8 | 21.7 |  |
| No declared preference (n=471) | 369 | 78.3 | 102 | 21.7 |  |
| Far left (n=145) | 99 | 68.3 | 46 | 31.7 |  |
| Left (n=98) | 66 | 67.3 | 32 | 32.7 |  |
| Green party (n=113) | 76 | 67.3 | 37 | 32.78 |  |
| Center (n=112) | 67 | 59.8 | 45 | 40.2 |  |
| Right (n=83) | 47 | 56.7 | 36 | 43.3 |  |
| Far right (n=221) | 143 | 64.7 | 78 | 35.3 |  |
| Science does people |  |  |  |  | 0.04 |
| more good than harm (n = 405) | 263 | 64.9 | 142 | 35.1 |  |
| about as much good as harm (n = 654) | 449 | 68.6 | 205 | 31.38 |  |
| more harm than good (n = 125) | 104 | 83.3 | 21 | 16.7 |  |

**Table 4:**
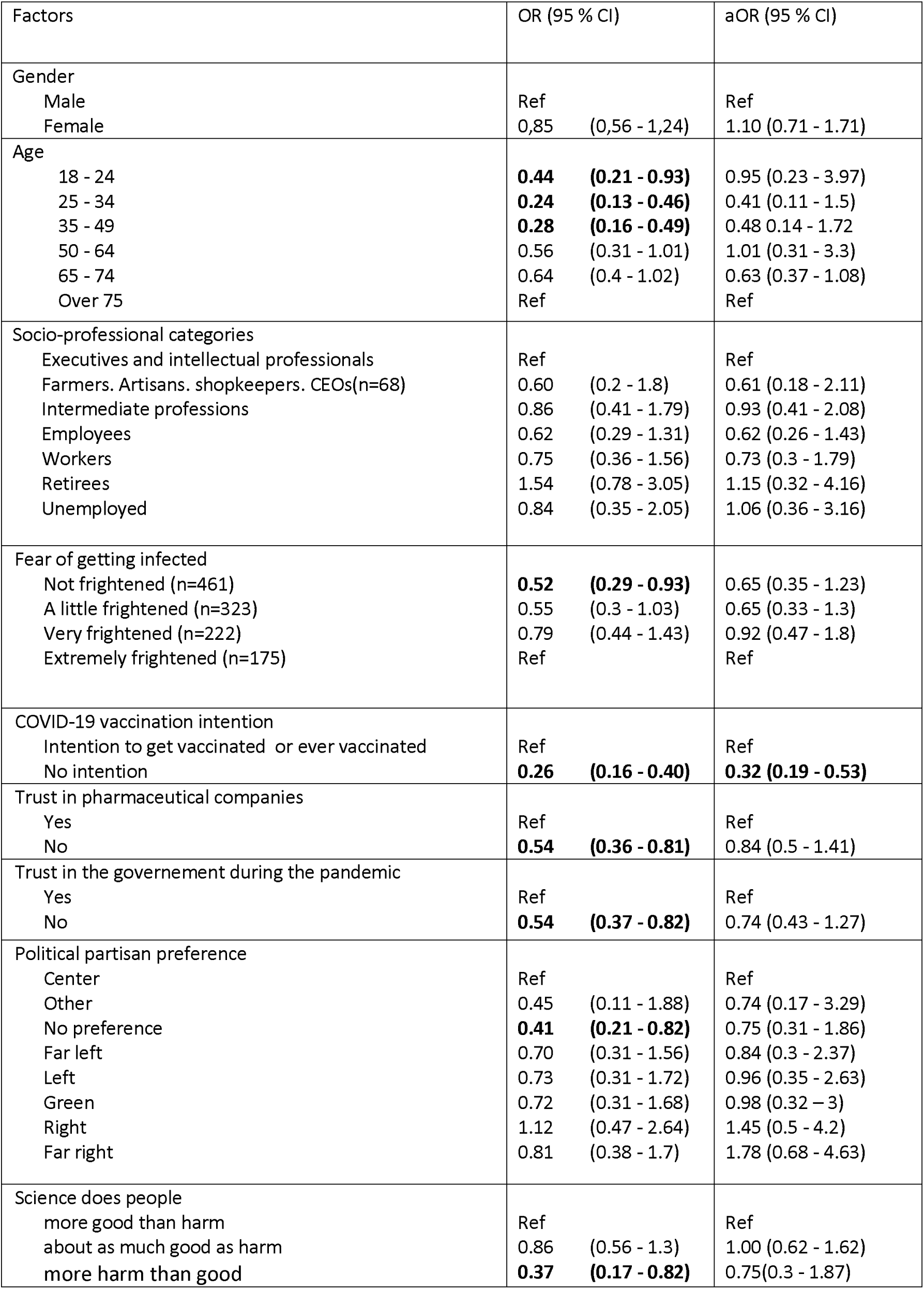
Factors associated with support for COVID-19 vaccine mandates for HCWs in opponent to a COVID-19 mandate for the general population in multivariable analysis (Ref: reference, variables with a p-value <0.05 are in bold OR: odds ratio, aOR: adjusted odds ratio, CEO: chief executive officer)

| Factors | OR (95 % CI) | aOR (95 % CI) |
| --- | --- | --- |
| Gender |  |  |
| Male | Ref | Ref |
| Female | 0.85 (0.56 - 1.24) | 1.10 (0.71 - 1.71) |
| Age |  |  |
| 18 - 24 | **0.44 (0.21 - 0.93)** | 0.95 (0.23 - 3.97) |
| 25 - 34 | **0.24 (0.13 - 0.46)** | 0.41 (0.11 - 1.5) |
| 35 - 49 | **0.28 (0.16 - 0.49)** | 0.48 (0.14 - 1.72) |
| 50 - 64 | 0.56 (0.31 - 1.01) | 1.01 (0.31 - 3.3) |
| 65 - 74 | 0.64 (0.4 - 1.02) | 0.63 (0.37 - 1.08) |
| Over 75 | Ref | Ref |
| Socio-professional categories |  |  |
| Executives and intellectual professionals | Ref | Ref |
| Farmers. Artisans. shopkeepers. CEOs(n=68) | 0.60 (0.2 - 1.8) | 0.61 (0.18 - 2.11) |
| Intermediate professions | 0.86 (0.41 - 1.79) | 0.93 (0.41 - 2.08) |
| Employees | 0.62 (0.29 - 1.31) | 0.62 (0.26 - 1.43) |
| Workers | 0.75 (0.36 - 1.56) | 0.73 (0.3 - 1.79) |
| Retirees | 1.54 (0.78 - 3.05) | 1.15 (0.32 - 4.16) |
| Unemployed | 0.84 (0.35 - 2.05) | 1.06 (0.36 - 3.16) |
| Fear of getting infected |  |  |
| Not frightened (n=461) | **0.52 (0.29 - 0.93)** | 0.65 (0.35 - 1.23) |
| A little frightened (n=323) | 0.55 (0.3 - 1.03) | 0.65 (0.33 - 1.3) |
| Very frightened (n=222) | 0.79 (0.44 - 1.43) | 0.92 (0.47 - 1.8) |
| Extremely frightened (n=175) | Ref | Ref |
| COVID-19 vaccination intention |  |  |
| Intention to get vaccinated or ever vaccinated | Ref | Ref |
| No intention | **0.26 (0.16 - 0.40)** | **0.32 (0.19 - 0.53)** |
| Trust in pharmaceutical companies |  |  |
| Yes | Ref | Ref |
| No | **0.54 (0.36 - 0.81)** | 0.84 (0.5 - 1.41) |
| Trust in the government during the pandemic |  |  |
| Yes | Ref | Ref |
| No | **0.54 (0.37 - 0.82)** | 0.74 (0.43 - 1.27) |
| Political partisan preference |  |  |
| Center | Ref | Ref |
| Other | 0.45 (0.11 - 1.88) | 0.74 (0.17 - 3.29) |
| No preference | **0.41 (0.21 - 0.82)** | 0.75 (0.31 - 1.86) |
| Far left | 0.70 (0.31 - 1.56) | 0.84 (0.3 - 2.37) |
| Left | 0.73 (0.31 - 1.72) | 0.96 (0.35 - 2.63) |
| Green | 0.72 (0.31 - 1.68) | 0.98 (0.32 - 3) |
| Right | 1.12 (0.47 - 2.64) | 1.45 (0.5 - 4.2) |
| Far right | 0.81 (0.38 - 1.7) | 1.78 (0.68 - 4.63) |
| Science does people |  |  |
| more good than harm | Ref | Ref |
| about as much good as harm | 0.86 (0.56 - 1.3) | 1.00 (0.62 - 1.62) |
| more harm than good | **0.37 (0.17 - 0.82)** | 0.75 (0.3 - 1.87) |

## Discussion

In this survey, we observed that the opinion of the general population on a mandatory COVID-19 vaccination policy was split, as 43 % of the respondents were in favor, 15 % were undecided and 41.9 % were opposed to it. Among the opponents to such a policy, around one third was in favor of a COVID-19 mandatory vaccine for HCWs.

France is known as a “vaccine-hesitant” country [9], and may be a reluctant country to a mandatory COVID-19 vaccination. The proportion of opponents to a mandatory COVID-19 vaccine in France is not far from the 51% proportion observed in a German study carried out in June and July 2020 [15]. We observed a higher proportion of opponents in France than in the USA, Greece respectively 17.3 % and 25.7 % [16,17] In Australia, 73% of the population said they would support the government requiring the coronavirus vaccine for activities such as travel, work, and study, and only 9 % were clearly opposed to a mandatory COVID-19 vaccine [18].

We observed that older age and very high fear of COVID-19 were associated with support of COVID-19 vaccine mandate for the general population. These factors were also identified in other European studies about COVID-19 vaccine mandates [15,17] were also associated with intentions to get vaccinated [19,20]. Intentions to get vaccinated or vaccinated status were highly associated with support for COVID-19 mandatory vaccination, it is not surprising to identify common determinants.

French reluctance to COVID-19 mandatory vaccination may in part be explained by some questions about mandatory vaccination. In December 2020, before the launch of the vaccine Campaign, the President of the French Republic promised that the vaccine would not be made mandatory. On the 12^th^ of July 2021, while the delta variant spread in France, he announced mandatory COVID-19 vaccination for HCWs and other exposed professions and the “COVID-19 passport” extension (Complete vaccine schedule, or COVID-19 infection in the past 6 months, or a negative SARS-Cov-2 test in the past 72 hours) for the general population to attend public settings (such as restaurants, movie theaters, shopping centers, etc…). Since this announcement, 13 million French people have received their first dose of vaccine, and vaccine coverage reached 85.1 % of the eligible population on the 8^th^ of September 2021. At the time this article is being written, the movement against “COVID-19 passport”, that protests every week, does not seem to be growing and is not supported by the majority of the French population. We observed that vaccinated or individuals who intend to get vaccinated could be opposed to COVID-19 vaccine mandates. In the United Kingdom, vaccine passports would make a large minority of individuals no more or less inclined to accept a COVID-19 vaccine, and individuals with definite intentions to get vaccinated were less inclined to get vaccinated if a vaccine passport was implemented [21]. It remains unclear if “COVID-19 passports” are more acceptable than COVID-19 mandatory vaccination for the general population. Indeed, mandatory COVID-19 vaccination for the general population is a highly politicized issue in the context of the 2022 presidential election campaign. We observed that lack of trust in the government during the pandemic and partisanship of far left and green parties were associated with a greater opposition to a mandatory COVID-19 vaccine policy. The influence of political identities on attitudes to vaccines was also previously observed for intention to get vaccinated against COVID-19 in France [12]. In the USA, Democrats, in Australia major party voters were more likely to be in favor of a mandatory COVID-19 vaccination than Republicans [16,18] In contrast, in Germany, political preferences do not seem to be associated with attitudes toward a mandatory COVID-19 vaccination [15]. It has been previously observed that attitudes toward vaccine mandates were even more influenced by partisan orientations than vaccination intentions [22]. In addition, since the 15th of October, COVID-19 tests in asymptomatic individuals to obtain “COVID-19 passports” are no longer be free in France. This appears to be a backdoor way of making vaccination almost compulsory.

A mandatory COVID-19 vaccination would lead to an increase in vaccine coverage as currently observed in French HCWs. HCWs COVID-19 vaccine coverage was 62.4% on the 12^th^ of July and reached 88.4% on the 6^th^ of September. However, a COVID-19 vaccine mandate might be counterproductive particularly if it is not acceptable for a great majority of the population [23]. Such a policy can have detrimental consequences: less uptake of other vaccines, decrease in adherence to personal protective measures, enhancement of suspicion of both vaccines in general and public health authorities, and reduction in autonomy in the decision making. A detrimental effect of a mandatory COVID-19 vaccine policy is quite uncertain in France. Santé Publique France has observed an increase in vaccine coverage of non-mandatory vaccines since the extension of mandatory vaccinations in infants, and a slight increase in the proportion of the French population favorable to vaccines in general [24]. After a period of reluctance, acceptability of COVID-19 mandatory vaccination will probably increase.. In the past, the rate of favorable opinions toward mandatory childhood vaccines increased after the extension of the number of mandatory vaccines in 2018 [24]. Furthermore, in July 2021 in an opinion poll, 58 % of the respondents were in favor of a COVID-19 vaccine mandate for all [25]. The COVID-19 passport could be considered as a form of COVID-19 vaccine mandate, a majority of the French general population (58 %) has a favorable opinion about the COVID-19 passport [26].

Our study suffers from several limitations. First, we can address the representativeness of participants in comparison with the French general population. Sample size is limited for the younger age groups, however the observations have been weighted for age, gender, professional categories, and living areas. Older age and antecedents or intention to take up COVID-19 vaccination were great predictors of attitudes toward COVID-19 vaccine mandates. The survey was an internet-based survey, individuals without access to technologies or disabilities are probably under-represented in our sample. In addition, undecided respondents were not asked about their attitudes to a mandatory COVID-19 vaccination for HCWs. As we observed that one third of the opponents to COVID-19 vaccine mandates in the general population were in favor of specific mandates for HCWs, we cannot estimate the true proportion of the population in favor of a mandatory COVID-19 vaccination for HCWs.

In conclusion, opinions toward COVID-19 vaccine mandates were split in France in May 2021. COVID-19 vaccine mandates is a highly political issue, in the context of the next French presidential election. Despite the implementation of the COVID-19 passport and COVID-19 vaccine mandates for HCWs and exposed professions, France seems to have hit the glass ceiling of COVID-19 vaccine coverage. In addition, disparities are observed between regions, and French overseas territories. If another wave hits France in the Autumn and if a more comprehensive outreach program is not put in place by then, the dilemma might well be: what would be less unacceptable: mandatory vaccination, or new containment measures?

## Data Availability

The data are available by request to the corresponding author.

## Acknowledgements

The authors would like to thank Cyril Bérenger (Database manager, ORS PACA), Sébastien Cortaredona (Statistician, IRD), Lisa Fressard (Statistician, ORS PACA), Gwenaelle Maradan (Logistician, ORS PACA) and Alvaro Sanchez (Statistician) for their data collection and analysis, Glyn Thoiron for English editing and Delphine Grison for her help.

## Conception or design of the work

PV, AGB, JKW, EBN, OL, MB

## Acquisition of the data

PV, JKW

## Analysis

PV, JKW, AGB

## Interpretation of data for the work

PV, OL, EBN, PPW, AGB

